# Automated detection of adult autism from vowel acoustics using machine learning

**DOI:** 10.64898/2026.04.03.26350102

**Authors:** Georgios P. Georgiou, Maria Paphiti

## Abstract

Autism spectrum disorder (ASD) is a neurodevelopmental condition for which timely and accurate detection remains a major clinical priority. Early and reliable identification is important because it can facilitate access to assessment, diagnosis, and appropriate support; however, current diagnostic pathways still rely largely on behavioural evaluation and clinical judgement. In this context, machine-learning (ML) approaches have attracted growing interest because they can identify subtle and complex patterns in speech data that may not be easily captured through conventional methods. The current study capitalizes on this potential by developing and evaluating ML models for distinguishing autistic individuals from neurotypical individuals based on speech features. More specifically, acoustic features of vowels, including fundamental frequency (F0), first three formants (F1, F2, F3), duration, jitter, shimmer, harmonics-to-noise ratio (HNR), and intensity, were elicited from 18 autistic adults and 18 neurotypical adults through a controlled production task. Then, four supervised ML models were trained and evaluated on these features: LightGBM, Random Forest, Support Vector Machine, and XGBoost. All models demonstrated good classification performance, with the best-performing model achieving a strong discriminability of 89%. The explainability analysis identified F0 as the most influential predictor by a substantial margin, followed by intensity, F3, and F1, while duration, shimmer, HNR, jitter, and F2 contributed more modestly. These findings demonstrate that vowel acoustics contain clinically relevant information for distinguishing autistic from neurotypical adult speech and highlight the potential of interpretable, speech-based ML as a transparent and scalable aid for ASD screening and assessment.

## 1. Introduction

Autism spectrum disorder (ASD) or simply autism is a lifelong neurodevelopmental condition characterized by persistent difficulties in social communication and social interaction, alongside restricted and repetitive patterns of behaviour, interests, or activities (Centers for Disease Control and Prevention, 2025). Speech and language differences are common in ASD, although they are heterogeneous across individuals and across development (Mody & Belliveau, 2013; Vogindroukas et al., 2022). These differences span a broad continuum, such as prosodic variation in pitch, intensity, and speech rate, as well as atypical language use, pragmatic difficulties, and challenges in reciprocal conversation (Hage et al., 2021; Klusek et al., 2014). These findings yield the importance of examining speech biomarkers in ASD, because speech constitutes a direct, observable, and objectively measurable signal of communication. Unlike higher-level language abilities, which are shaped by cognitive, contextual, and sociocultural factors (Swineford et al., 2014), speech can be quantified through acoustic features such as pitch, timing, voice quality, and articulation (Ng et al., 2026). This makes speech particularly suitable for systematic analysis and for identifying reliable, low-level markers of atypical communication that are less dependent on linguistic content or task demands.

The growing global burden of ASD has intensified interest in artificial intelligence (AI) and machine learning (ML) as tools for earlier and more objective detection (Santomauro et al., 2025). Current diagnostic practice still relies primarily on developmental history, behavioural observation, and clinician judgement, while no established biological biomarkers are in routine clinical use, making timely identification challenging (Cortese et al., 2025). In this context, ML has emerged as a promising approach because it can analyse large, complex, and multimodal datasets and detect patterns not easily captured through conventional assessment alone (Solek et al., 2025). Recent research has increasingly integrated ML with behavioural, neuroimaging, genetic, and eye-tracking data to support earlier ASD identification (e.g., Frye et al., 2019). Early identification is important because it enables timely intervention, which has been associated with improvements in social communication, cognition, and related developmental outcomes (Collins et al., 2025; Fuller & Kaiser, 2020). Among candidate biomarkers under investigation, speech has gained growing attention as a potentially informative marker of neurodevelopmental conditions, including autism, and ML offers a promising means of detecting subtle speech patterns that may support ASD identification.

### 1.1 Acoustic differences between autistic and neurotypical speakers

Autistic and neurotypical speech can differ across several acoustic dimensions, including prosodic, spectral, temporal, voice-quality/phonatory, and amplitude-related measures. Among these, differences in fundamental frequency (F0), the main acoustic correlate of perceived pitch, are among the most robust findings in the ASD speech literature. A systematic review and meta-analysis by Asghari et al. (2021) concluded that mean pitch and pitch range reliably distinguish autistic from neurotypical speakers, with pitch-range effects being particularly consistent across studies. Bonneh et al. (2011) found that autistic children showed a significantly larger pitch range and greater pitch variability over time than controls, challenging the traditional view that autistic speech is characteristically monotone. More recent work has also reported higher F0 in autistic children, although not always a wider F0 range, suggesting that pitch atypicality may manifest differently depending on the speech sample and analytic approach (Beccaria et al., 2025).

Formant-related evidence is more limited but important because it may reflect differences in articulatory configuration and vocal-tract resonance. Briend et al. (2023) reported that autistic children showed lower average F1 than neurotypical children, and that F1 was among the strongest discriminators of ASD relative to both neurotypical and heterogeneous control groups. Similarly, Sadiq et al. (2025) found that autistic children had significantly lower F1 and F2, while F3 showed a near-significant trend. In adults, however, findings point to somewhat different patterns. Kissine et al. (2021) reported greater articulatory stability in autistic adults’ native vowel production, based on F1, F2, and F3 measures. Related summaries of this work suggest smaller F1–F3 dispersion in autistic adults, interpreted as more stable but less flexible articulatory patterns. At the same time, not all studies report formant differences. For example, Maffei et al. (2025) found no significant differences in F2-based vowel measures between verbal autistic and non-autistic children, although differences were more apparent in minimally verbal autistic children. Moreover, temporal differences are also well documented. Asghari et al. (2021) found that voice duration was significantly longer in autistic speakers overall, identifying duration as another cross-study marker. In a study of verbal autistic preschoolers, Maes et al. (2024) likewise reported slightly exaggerated syllable duration, even though the overall acoustic structure of autistic and neurotypical speech was largely similar.

Evidence for voice-quality measures such as jitter, shimmer, and harmonics-to-noise ratio (HNR) is more mixed and may vary by age. In children, Bone et al. (2014) reported that higher jitter contributed to perceived overall voice severity in autistic speech, while jitter- and HNR-related measures were associated with ASD severity in spontaneous speech. Briend et al. (2023) also found that the acoustic profile most predictive of ASD included higher shimmer and lower jitter skewness, with mean shimmer emerging as one of the strongest discriminators. In adults, however, the pattern may differ. Kissine and Geelhand (2019) found lower jitter and shimmer in autistic adults than in matched neurotypical adults, suggesting greater phonatory stability rather than instability. By contrast, Drimalla et al. (2020) found no significant group differences in jitter or shimmer in a standardized dialogue task. HNR also appears to have some discriminative potential. Briend et al. (2023) reported higher HNR in autistic children, interpreted as less noisy and more harmonic vocal output, and Bone et al. (2014) likewise linked HNR-related measures to ASD severity. Intensity, by contrast, has been less consistent as a marker. According to Asghari et al. (2021), intensity has not shown reliable case-control differences across studies. This was echoed by Sadiq et al. (2025), who found no significant intensity differences between autistic and neurotypical children despite detecting significant formant differences. However, Hubbard et al. (2017) reported greater intensity in adult males with ASD across emotional speech conditions.

Overall, previous research suggests that autistic and neurotypical speakers may differ across several acoustic aspects of speech. The clearest findings concern pitch-related measures and duration, while evidence for formants, jitter, shimmer, HNR, and intensity is more variable. Thus, the literature supports the view that speech in ASD can show measurable acoustic atypicalities, but these are not always uniform across studies.

### 1.2 ML classification of autistic and neurotypical speech

A second line of research has examined whether acoustic speech features can be used to classify autistic and neurotypical speakers through ML. Rather than focusing on single acoustic measures in isolation, these studies train models on combinations of prosodic, spectral, formant, and voice-quality variables. Some rely on classical acoustic parameters, while others use broader clinical feature sets or higher-dimensional spectral representations.

One of the clearest examples based on classical acoustic measures is Briend et al. (2023). Using speech from a nonword repetition task in children, the authors extracted 19 acoustic variables and applied receiver operating curve (ROC)-supervised k-means clustering with Monte Carlo cross-validation. Their model achieved 91% accuracy in distinguishing autistic from neurotypical children and 85% accuracy in distinguishing autistic children from a heterogeneous non-autistic comparison group. The most discriminative features were mean F1, mean HNR, mean shimmer, and jitter skewness, with sensitivity = 89% and specificity = 94%. In a related study, Guo et al. (2022) examined Mandarin-speaking children during a lexical tone picture-naming task. Their feature set included F0, F0 range, HNR, jitter, and shimmer, along with spectral voice-quality measures, and a Random Forest classifier achieved 78.5% accuracy, with shimmer and jitter emerging as especially robust features.

Beccaria et al. (2022) extracted clinically oriented acoustic variables, including F0, F1, F2, F3, jitter, HNR, and related spectral-energy measures, from Italian-speaking children with ASD and tested several supervised classifiers. Their best models reached 83% accuracy on the held-out test set, while Decision Trees and Support Vector Machines (SVMs) achieved area under the curve (AUC) values of 88%. The SVM model also reached 100% recall, 67% precision, and 80% F1. Mohanta et al. (2022), focusing on English vowels produced by Indian children with ASD and neurotypical development, used features including F0, F1–F5, energy, and additional source-filter measures. Comparing several models, they reported classification accuracy up to 98.17%, with the best result obtained by a Probabilistic Neural Network. Other studies suggest that speech-based classification may be possible even very early in development. Pokorny et al. (2017) examined pre-linguistic vocalisations from 10-month-old infants later diagnosed with ASD and matched controls. Both a linear-kernel SVM and a one-layer bidirectional neural network achieved 75% subject-wise accuracy in a subject-independent three-fold cross-validation design.

The ML literature suggests that speech acoustics contain diagnostically useful information for distinguishing autistic from neurotypical speakers, especially in child samples and in structured elicitation tasks such as nonword repetition, vowel production, or lexical tone production. Across studies, informative variables often come from a combination of pitch-related, formant-based, and voice-quality domains. However, reported performance depends strongly on recording context, sample size, and validation strategy. Accordingly, current findings support the promise of speech-based ML as a potential diagnostic aid rather than a standalone diagnostic tool, and they suggest that future work should combine interpretable acoustic biomarkers with stronger cross-context validation procedures.

### 1.3 The current study

Despite growing interest in speech-based markers of autism, important gaps remain in the literature. First, much of the existing research has focused on children, whereas relatively less is known about the acoustic characteristics of autistic adults. Second, many ML studies have relied on English or other widely studied languages, with very limited evidence from understudied linguistic varieties. Third, previous work has often drawn on broader or less interpretable feature sets, including high-dimensional spectral representations or deep-learning approaches, which may achieve good classification performance but make it more difficult to identify which specific speech characteristics drive model decisions. Finally, although several studies report promising classification results, many are based on small and highly controlled datasets, and fewer studies combine strong predictive modelling with explainable artificial intelligence (XAI) methods that can clarify the relative contribution of individual acoustic features.

The present study bridges these gaps by developing an ML-based approach for distinguishing adults with autism from neurotypical adults based on vowel production. The following features were elicited through a controlled production task from adult speakers of Cypriot Greek: F0, F1, F2, F3, duration, jitter, shimmer, HNR, and intensity. Rather than limiting the analysis to descriptive group comparisons, the study evaluates the extent to which these acoustic features can be integrated within supervised ML models to support the accurate classification of autistic versus neurotypical speech. The study is original in several respects. To our knowledge, it is the first to develop and test an ML-based approach for autism detection in speakers of Greek, a linguistic and cultural context that remains largely absent from the literature. It is also novel in its use of a theory-driven set of classical acoustic variables, rather than relying mainly on less interpretable high-dimensional representations. In addition, the study incorporates XAI methods to identify the acoustic features that contribute most strongly to model predictions, thereby enhancing the interpretability and clinical relevance of the proposed ML approach.

## 2. Methodology

### 2.1 Participants

Thirty-six native speakers of Cypriot Greek participated in the study, including 18 individuals with ASD (15 males/3 females) aged between 18 and 40 years (*M*_age_ = 24.1 years; *SD* = 6.43) and 18 participants with neurotypical development (ND) (15 males/3 females) aged 18 to 42 years (*M*_age_ = 24.5 years; *SD* = 7.11). Participants in the ASD group had an officially documented clinical diagnosis established by experienced clinicians, neurologists, and psychiatrists in accordance with the Diagnostic and Statistical Manual of Mental Disorders, Fifth Edition (DSM-5) criteria. They were recruited from specialized autism centers and secondary schools in Cyprus. Individuals with a history of intellectual disability or co-occurring language disorder were excluded. The ND participants were recruited across Cyprus and had no reported history of neurodevelopmental, language, cognitive, or behavioral disorders.

The two groups were comparable on several background variables. There were no significant differences in chronological age, *t*(34) = -0.17, *p* = 0.86, nonverbal IQ (Raven et al., 2018), *t*(34) = -1.97, *p* = 0.06, educational attainment (UNESCO Institute for Statistics, 2012), *W* = 149, *p* = 0.68, or gender distribution, χ²(1) = 0, *p* = 1.00. The groups also did not differ significantly on verbal fluency performance (Kosmidis et al., 2004): semantic fluency task: mean word production, *t*(34) = -1.65, *p* = 0.11; mean cluster size, *t*(34) = 0.46, *p* = 0.65, number of switches, *t*(34) = -1.72, *p* = 0.09; phonemic fluency task, mean word production, *t*(34) = -1.97, *p* = 0.06; mean cluster size, *t*(34) = -0.83, *p* = 0.41; number of switches, *t*(34) = -0.83, *p* = 0.08.

As expected, the groups differed significantly on measures related to autistic traits, based on the Greek versions of the Empathy Quotient (EQ) (Pehlivanidis et al., 2021) and the Autism-Spectrum Quotient (AQ) (Baron-Cohen et al., 2001) self-report questionnaires. The ASD group had lower EQ scores, with a mean of 29.7 (*SD* = 12.4), compared with 43.1 (*SD* = 10.2) in the ND group, *t*(34) = -3.55, *p* = 0.001. Conversely, the ASD group had higher (AQ) scores, with a mean of 29.6 (*SD* = 6.49), compared with 14.8 (*SD* = 5.96) in the ND group, *t*(34) = 7.11, *p* < 0.001. All participants reported normal or corrected-to-normal vision and hearing. Prior to participation, all individuals, and guardians where applicable, received written information about the study procedures and their rights and provided informed consent in accordance with the Declaration of Helsinki.

### 2.2 Materials

The stimulus set comprised disyllabic pseudowords presented in four configurations: / sVsa/, /sV□sa/, /□Vsa/, and /V□sa/, where V represented the target vowel. In each condition, the first syllable contained one of the five Greek vowels /i e a o u/. All items were designed to follow Greek phonotactic rules and to sound word-like, while remaining devoid of semantic meaning.

### 2.3 Procedure

#### 2.3.1 Production Test

Each participant completed the production task individually in a quiet testing environment. During the session, participants were seated at a desk facing a computer monitor at a distance of approximately 50–70 cm. The stimuli were displayed in Standard Modern Greek orthography, and participants were asked to read each item aloud using a natural speaking rate and comfortable vocal intensity. Speech was recorded in WAV format with a Zoom H5 digital recorder at a sampling rate of 44.1 kHz and 16-bit resolution. To ensure consistency across speakers, both recorder placement and mouth-to-recorder distance were kept constant, and recording levels were calibrated before each session to prevent clipping. Each participant produced a total of 80 words, corresponding to five vowels across four phonetic contexts with four repetitions each, yielding 2,880 words across the full sample. Stimuli were randomized within each repetition block. Before the main task, participants completed a short familiarization phase to ensure correct reading of the items; these practice productions were excluded from the analyses. Tokens containing reading errors, disfluencies, or excessive background noise were re-recorded whenever possible; otherwise, they were excluded. Short breaks of up to five minutes were permitted between repetition blocks.

#### 2.3.2 Feature Extraction

The target words were segmented and analyzed in Praat (Boersma & Weenink, 2026). Visual inspection of spectrograms and waveforms was used to identify relevant acoustic landmarks, allowing the delimitation of vowel boundaries and the extraction of acoustic measures including F0, F1, F2, F3, duration, jitter, shimmer, HNR, and intensity. Analyses were conducted using a window length of 0.025 s, pre-emphasis set at 50 Hz, and a spectrogram display ceiling of 5500 Hz (see Georgiou & Savva, 2025). Measurements were taken over the vowel interval for each token. In the /□sVsa/ and /sV□sa/ contexts, the measurement interval began at the end of the initial /s/ frication, corresponding to vowel onset, and ended at the offset of vocalic periodicity immediately before the following consonant. In the /□Vsa/ and /V□sa/ contexts, where no initial fricative was present, the interval extended from the onset of vocalic periodicity to its offset before the /s/. For F0 extraction, the pitch range was set to 75–300 Hz. F0, F1, F2, F3, and intensity were measured at the temporal midpoint of the vowel to reduce coarticulatory influence from surrounding segments. Vowel duration was defined as the full temporal extent of the vowel. Jitter, shimmer, and HNR were derived using Praat’s jitter (local), shimmer (local), and harmonicity (cc) functions, respectively.

#### 2.3.3 ML modelling and evaluation

Four supervised ML algorithms were implemented to classify ASD and ND participants: LightGBM, Random Forest, SVM, and XGBoost. These models were chosen because they represent complementary and well-validated approaches to binary classification on tabular data. Random Forest provides a robust ensemble baseline based on bagging, SVM offers a margin-based classifier for two-class separation, and XGBoost and LightGBM represent high-performing gradient-boosted tree frameworks capable of modelling complex non-linear relations and feature interactions (Breiman, 2001; Chen & Guestrin, 2016; Cortes & Vapnik, 1995; Ke et al., 2017). All analyses were conducted in R (R Core Team, 2026) using the *caret* framework for model training and evaluation.

The dataset included acoustic features (F0, F1, F2, F3, duration, jitter, shimmer, HNR, and intensity) as predictors and group membership (ASD vs ND) as the outcome variable. The dataset was then randomly partitioned into training (80%) and testing (20%) subsets using stratified sampling to preserve class proportions. Feature scaling was applied to all predictors using centering and standardization based on the training data, with the same transformation applied to the test set.

Hyperparameter tuning was performed separately for each model using cross-validation procedures aimed at maximizing AUC. LightGBM was trained using gradient boosting decision trees with predefined hyperparameters, including learning rate, number of leaves, maximum tree depth, feature subsampling, and regularization terms. Model complexity was controlled via 5-fold cross-validation with early stopping, where training was halted if performance did not improve for a specified number of iterations. The optimal number of boosting rounds was selected based on cross-validated AUC. Random Forest tuning was conducted within the caret framework using 5-fold cross-validation. A grid search over the number of variables randomly sampled at each split (mtry) was performed, while the number of trees was fixed at 500. SVM with a radial basis function kernel was tuned using 5-fold cross-validation. The model automatically optimized key hyperparameters, including the cost parameter (C) and kernel width (σ), through an internal grid search defined by the tuning length. XGBoost was similarly tuned using 5-fold cross-validation within caret. A grid search was applied over multiple hyperparameters, including the number of boosting rounds, learning rate, maximum tree depth, subsampling ratio, and column sampling parameters, with model selection based on cross-validated AUC. Model performance was evaluated using accuracy, precision, recall (sensitivity), F1-score, and AUC (see Georgiou & Theodorou, 2024).

To improve model interpretability, an XAI analysis was conducted using Shapley Additive Explanations (SHAP) values (Lundberg & Lee, 2017). SHAP values were calculated for the predictor variables to quantify the contribution of each feature to individual model predictions relative to a baseline value. These values were then examined using the *shapviz* package in R. This approach was used to provide both local and global interpretability of the model.

## 3. Results

Overall, the models demonstrated good ability to distinguish between individuals with ASD and ND. Among the evaluated approaches, the ensemble tree-based models (LightGBM, Random Forest, and XGBoost) showed similarly strong performance, with Random Forest demonstrating a slight advantage, suggesting that these models were effective in capturing patterns that distinguished between ASD and ND. In contrast, SVM demonstrated comparatively lower performance, suggesting reduced effectiveness relative to the ensemble-based methods. Table 1 presents the evaluation metrics of all four ML models. Figure 1 illustrates the confusion matrices for the four ML models used to classify ASD and ND.

**Figure 1:**
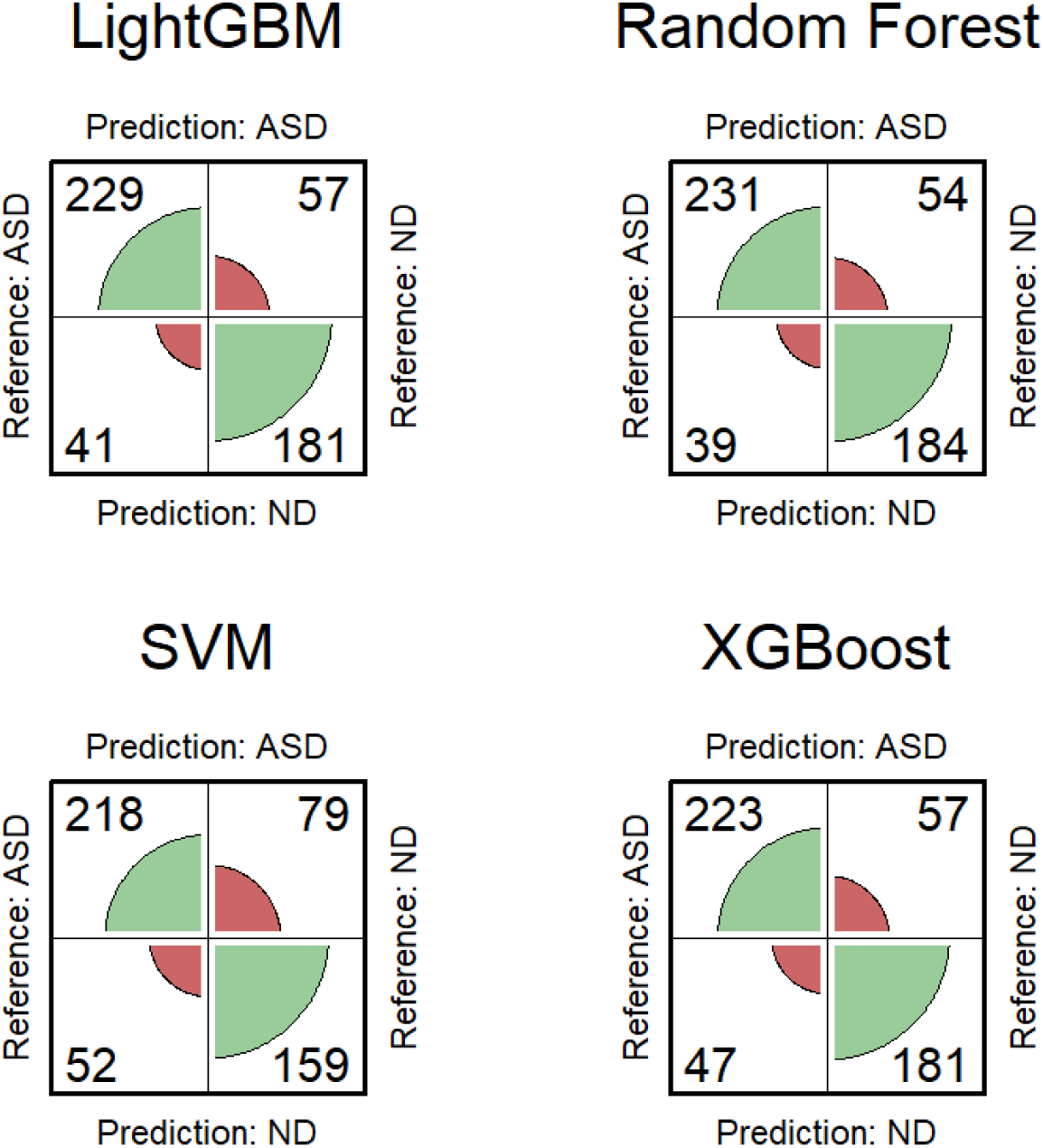
Confusion matrices for LightGBM, Random Forest, SVM, and XGBoost in ASD versus ND classification, illustrating the distribution of true positives (upper left), false positives (upper right), false negatives (lower left), and true negatives (lower right) for each model.

**Table 1:** Comparative performance of ML models for ASD versus ND classification

| Model | Metric |  |  |  |  |
| --- | --- | --- | --- | --- | --- |
|  | Accuracy | Precision | Recall | F1-score | AUC |
| LightGBM | 0.807 | 0.801 | 0.848 | 0.824 | 0.886 |
| Random Forest | 0.817 | 0.811 | 0.856 | 0.832 | 0.892 |
| SVM | 0.742 | 0.734 | 0.807 | 0.769 | 0.812 |
| XGBoost | 0.795 | 0.796 | 0.826 | 0.811 | 0.874 |

As seen in Figure 2, the ROC curves illustrate the classification performance of the four ML models in distinguishing between ASD and ND. Overall, all models yielded good discriminative ability, as indicated by curves that remained well above the diagonal reference line representing chance-level performance (AUC > 0.81). Among the models, Random Forest showed the strongest overall performance, with the highest AUC, closely followed by LightGBM and XGBoost, which also exhibited strong and comparable discriminative capacity. SVM demonstrated comparatively lower performance, with a ROC curve that lay closer to the diagonal line across much of the range, indicating reduced classification effectiveness relative to the ensemble-based methods.

**Figure 2.**
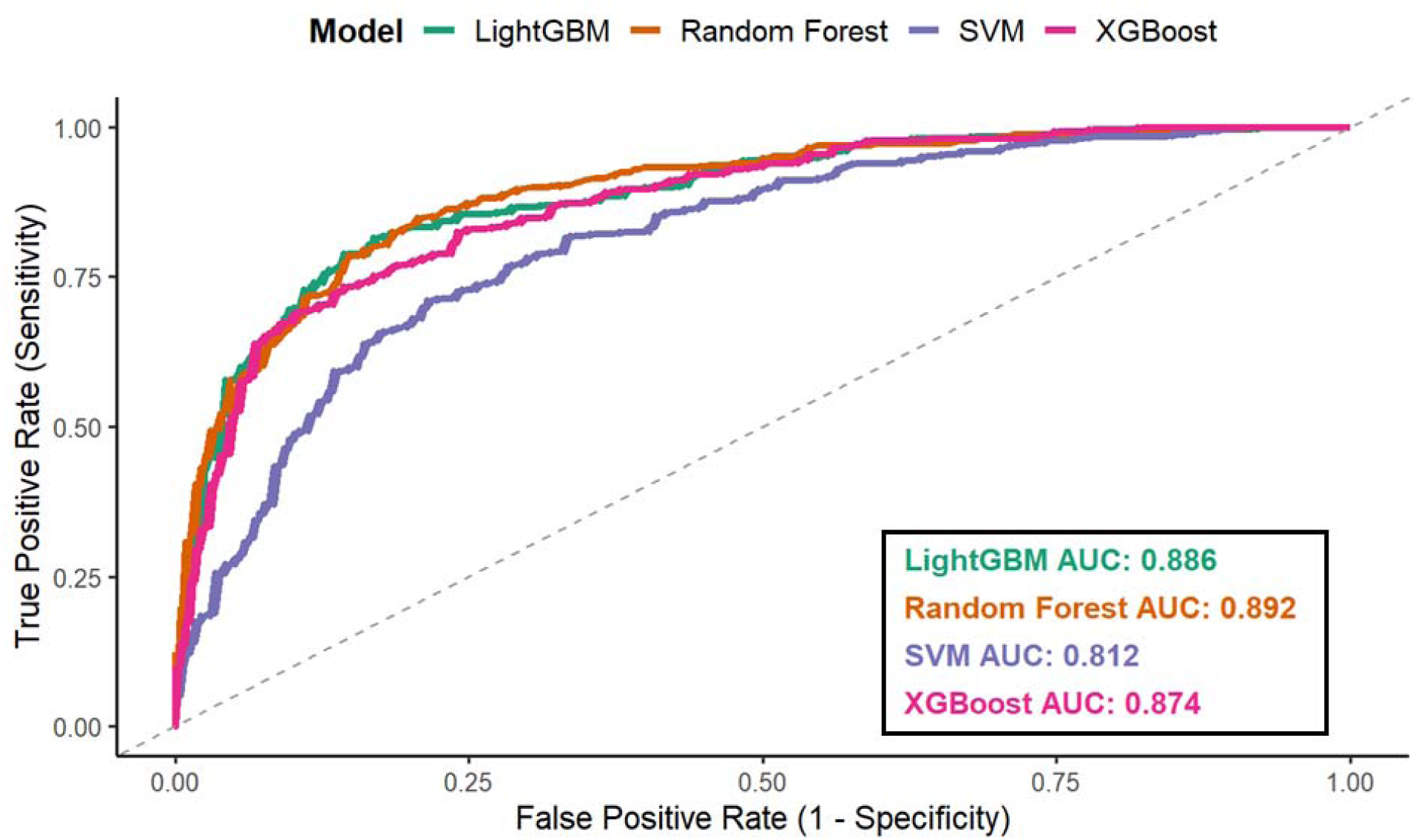
ROC curves and AUC values for ML models classifying ASD versus ND

SHAP was chosen for model interpretability due to its solid theoretical foundation in game theory, its ability to provide both local and global explanations, and its suitability for interpreting complex, non-linear models such as gradient boosting frameworks. LightGBM was selected for the explainability analysis because it was among the best-performing models in the study and because it allows efficient and exact computation of SHAP values through TreeSHAP. Although SHAP values can also be obtained for other tree-based models, such as Random Forest, the implementation is generally less direct and may be computationally less efficient.

The SHAP analysis indicated that F0 was the most influential feature in predicting ASD, showing substantially greater importance than all other variables (see Figure 3). Intensity emerged as the second most important feature, followed by F3 and F1, which contributed moderately to the model’s predictions. The remaining features – duration, shimmer, HNR, jitter, and F2 – showed comparatively lower but still meaningful contributions. The SHAP value distributions further illustrated that higher and lower values of these features exert differential effects on the prediction outcome, with several features demonstrating both positive and negative contributions depending on their value range. Overall, the results highlighted that a small subset of acoustic features, particularly F0 and intensity, drove most of the model’s predictive performance, while the remaining features played a more secondary role. Table 2 shows the mean absolute SHAP values for each acoustic feature, with bootstrap 95% confidence intervals (CIs). F0 was clearly separated from all others; intensity was also clearly separated from the lower group. There was a cluster of overlapping CIs among F3, F1, duration, shimmer, HNR, jitter, and F2, especially in the mid-lower range.

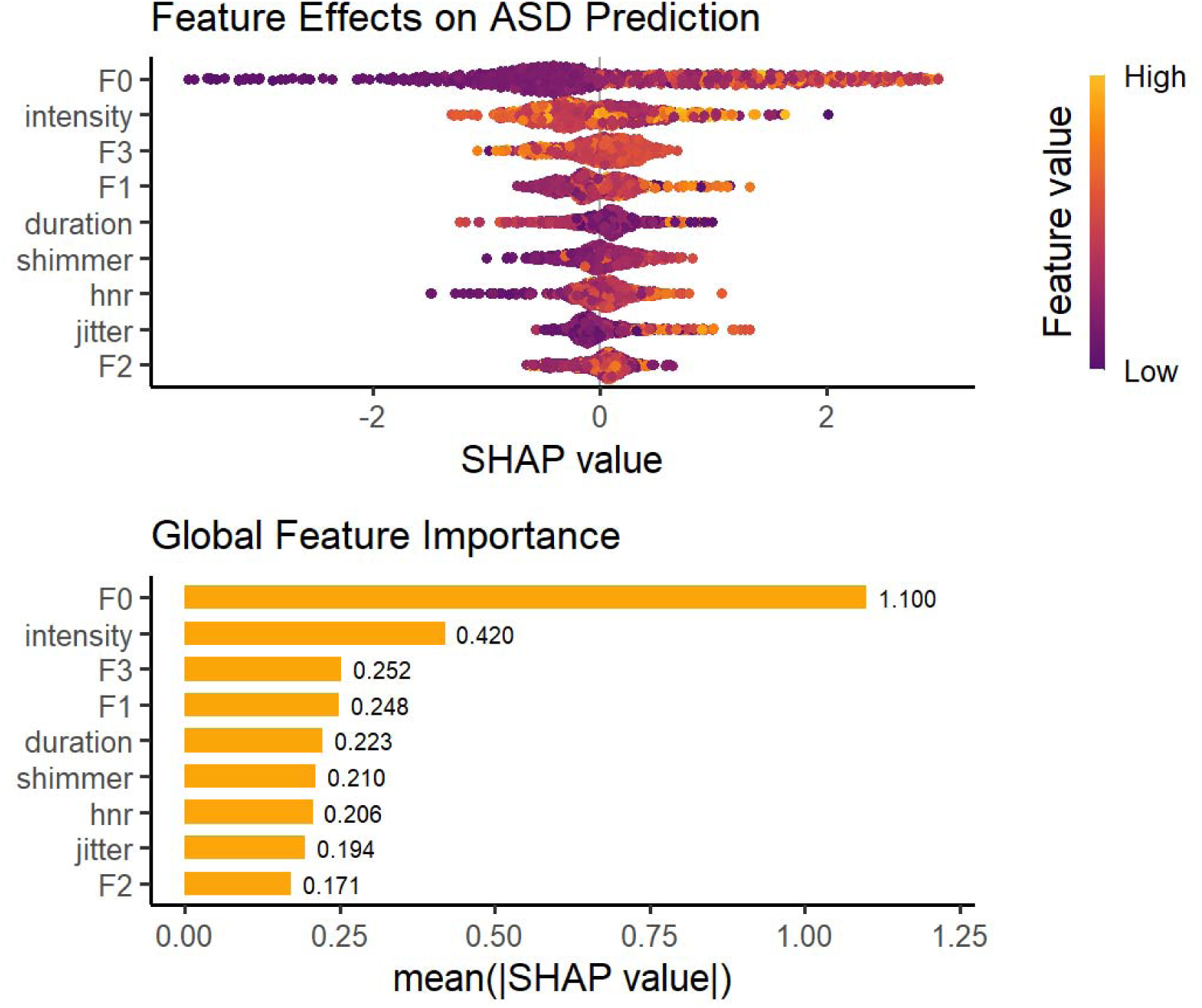

**Table 2:** Mean absolute SHAP values for each acoustic feature, with bootstrap 95% CIs (2.5th–97.5th percentiles; B = 2000). Higher values indicate greater average contribution to the model prediction (magnitude only; not direction).

| Feature | Mean Absolute SHAP | Bootstrap 95% CIs |
| --- | --- | --- |
| F0 | 1.100 | 1.03–1.171 |
| intensity | 0.420 | 0.393–0.447 |
| F3 | 0.252 | 0.236–0.269 |
| F1 | 0.248 | 0.232–0.268 |
| duration | 0.223 | 0.204–0.242 |
| shimmer | 0.210 | 0.194–0.227 |
| HNR | 0.206 | 0.187–0.227 |
| jitter | 0.194 | 0.177–0.212 |
| F2 | 0.171 | 0.159–0.184 |

## Discussion

The present study developed and evaluated an ML model for ASD detection based on vowel features in autistic and neurotypical adult speech. Overall, the findings indicated that vowel acoustics contain substantial information relevant to ASD classification in adulthood, since, across ML models, especially Random Forest, classification performance was consistently high. These findings extend previous work showing that speech measures can discriminate autistic from non-autistic speakers (Briend et al., 2023; Guo et al., 2022; Mohanta & Mittal, 2022), while also demonstrating that this potential is not restricted to children or to widely studied languages. In that respect, the present study contributes novel evidence from an adult Cypriot Greek sample and supports the broader view that speech may function as a promising, language-sensitive but still measurable biomarker domain in autism research.

A central finding of the study is that F0 emerged as the most influential predictor in the SHAP analysis, clearly exceeding all other variables in mean contribution to model output. This result is highly consistent with the literature identifying pitch-related measures as among the most robust acoustic correlates of autistic speech (Asghari et al., 2021). Although autism has often been stereotypically associated with monotone speech, empirical evidence has increasingly shown a more complex pattern, including elevated pitch, greater pitch variability, and atypical pitch modulation (Beccaria et al., 2025; Bonneh et al., 2011). Our findings reinforce the idea that pitch is not merely descriptively atypical in autism, but also diagnostically informative when incorporated into predictive models. Importantly, because the present study focused on adults, the results suggest that pitch-related atypicalities may remain detectable beyond childhood, even when speech and language abilities are otherwise relatively preserved.

A second important result is the contribution of intensity, which ranked just below F0 in the explainability analysis. Intensity has shown less consistency than pitch in prior case-control studies, with some reviews suggesting weak or unreliable group differences overall (Asghari et al., 2021), yet individual studies have reported elevated intensity under certain speaking conditions (Hubbard et al., 2017). The present findings suggest that even when intensity is not always a stable standalone group marker, it may still contribute meaningfully in combination with other acoustic cues. This highlights an important methodological point, as features that appear modest in univariate comparisons may become informative when modelled jointly, because ML algorithms can exploit multivariate interactions and nonlinear dependencies (Breiman, 2001; Chen & Guestrin, 2016). The moderate contributions of F1 and F3 also align with emerging evidence that articulatory-resonance properties may distinguish autistic and neurotypical speech, although less consistently than prosodic measures. Studies in children have reported lower F1 and, in some cases, lower F2 in autistic groups (Briend et al., 2023; Sadiq et al., 2025), while work on autistic adults has suggested greater articulatory stability and reduced flexibility in vowel production (Kissine et al., 2021; Kissine & Geelhand, 2019). Our results fit this broader pattern by indicating that formant-based information contributes to classification, but not as strongly as pitch. At the same time, the findings of this study suggest that F3, F1, duration, shimmer, HNR, jitter, and F2 form a secondary cluster of partially overlapping predictive cues rather than sharply differentiated markers. This pattern is theoretically plausible given the heterogeneity of ASD and the likelihood that no single acoustic feature fully captures the speech phenotype (Probol & Mieskes, 2024; Vogindroukas et al., 2022).

Our study highlights the clinical value of ML classifiers in ASD detection, because speech-based ML showed good discrimination between autistic and neurotypical adults. This is important for clinical practice because ASD diagnosis still relies heavily on expert behavioural assessment, which can be time-intensive and difficult to access promptly (Cortese et al., 2025). Recent reviews suggest that AI-based tools may help address this challenge by converting subtle communicative patterns into objective markers that can support earlier triage and referral decisions. More specifically, voice biomarkers have been identified as one of the most promising domains in AI-based autism detection (Rakotomanana & Rouhafzay, 2025), while explainability has been reported as essential if such systems are to be trusted and used responsibly in clinical contexts (Zhang et al., 2026). Together, these points strengthen the significance of the present study: ML classifiers are valuable not only because they can detect ASD-related speech patterns, but also because, when paired with explainable methods, they can provide transparent and clinically meaningful support for specialist judgement rather than replace formal diagnosis (Mohammadi et al., 2025).

## 5. Conclusions

Overall, the present study provides evidence that interpretable acoustic features can support reliable ML-based discrimination of autistic and neurotypical adult speech. The findings strengthen the case for speech as a scalable, non-invasive source of digital biomarkers in autism, which can assist the development of systems for ASD screening. These findings should, however, be interpreted in light of several limitations. The sample was relatively small and included a highly controlled reading task using pseudowords. Such a design was appropriate for isolating low-level acoustic properties, but it necessarily reduces ecological validity. Prior work has noted that strong performance in controlled speech datasets may not generalize straightforwardly to naturalistic or home-recorded settings, where linguistic variability, background noise, and speaker state introduce additional complexity (Robin et al., 2020). Future work should therefore test whether the current feature set retains predictive value in more spontaneous speech, across tasks, and in larger independent datasets. It will also be important to examine whether similar patterns hold across different Greek varieties and across more gender-balanced samples.

## Data availability

Data are available on reasonable request from the first author.

## Funding

This study forms part of the research project CULTURE/AWARD-YR/0523, awarded to the first author, Georgios P. Georgiou, by the Cyprus Research and Innovation Foundation.

## Conflict of Interest

The authors declare no conflicts of interest.

## Ethics Approval

The study was approved by the Cyprus National Bioethics Committee (approval no. EEBK EΠ 2024.01.57).

## Consent

All participants were informed about the aims of the study prior to participation, and written informed consent was obtained from the participants themselves and/or their legal guardians, in accordance with the Declaration of Helsinki.

## Acknowledgments

We sincerely thank all participants for taking part in this study. We also thank the Day Care Centre B, Pancyprian Association for Autism, for its assistance with participant recruitment and the Department of Secondary General Education of the Cyprus Ministry of Education, Sport, and Youth for granting permission (no. 07.30.002.002) to access upper secondary schools for data collection. This work was also supported by the Phonetic Lab at the University of Nicosia.

## Notes

### Competing Interest Statement

The authors have declared no competing interest.

